# SNPred outperforms other ensemble-based SNV pathogenicity predictors and elucidates the challenges of using ClinVar for evaluation of variant classification quality

**DOI:** 10.1101/2023.09.07.23295192

**Authors:** Ivan Molotkov, Daniel C. Koboldt, Mykyta Artomov

## Abstract

**Background:** Current single nucleotide variants (SNVs) pathogenicity prediction tools assess various properties of genetic variants and provide a likelihood of causing a disease. This information aids in variant prioritization – the process of narrowing down the list of potential pathogenic variants, and, therefore, facilitating diagnostics. Assessing the effectiveness of SNV pathogenicity tools using ClinVar data is a widely adopted practice. Our findings demonstrate that this conventional method tends to overstate performance estimates.

**Methods:** We introduce SNPred, an ensemble model specifically designed for predicting the pathogenicity of nonsynonymous single nucleotide variants (nsSNVs). To evaluate its performance, we conducted assessments using six distinct validation datasets derived from ClinVar and *BRCA1* Saturation Genome Editing (SGE) data.

**Results:** Across all validation scenarios, SNPred consistently outperformed other state-of-the-art tools, particularly in the case of rare and cancer-related variants, as well as variants that are classified with low confidence by most *in silico* tools. To ensure convenience, we provide precalculated scores for all possible nsSNVs.

We proved that the exceptionally high accuracy scores of the best models achieved for ClinVar variants are only attainable if the models learn to replicate misclassifications found in ClinVar. Additionally, we conducted a comparison of predictor performance on two distinct sets of BRCA1 variants that did not overlap: one sourced from ClinVar and the other from the SGE study. Across all *in silico* predictors, we observed a significant trend where ClinVar variants were classified with notably higher accuracy.

**Conclusions:** We provide a powerful variant pathogenicity predictor that enhances the quality of clinical variant interpretation and highlights important challenges of using ClinVar for SNV pathogenicity predictors evaluation.

## Introduction

With the availability of low-cost whole genome sequencing, it is now possible to comprehensively investigate the genetic variation in a patient in a single experiment. Yet every individual harbors thousands of sequence variants in the coding DNA region, only a fraction of which are expected to be deleterious. Identifying the small proportion of variants that may contribute to disease risk remains a significant challenge in human genetics. While it is not practically feasible at this time to measure the functional impact of all variants found in a particular patient experimentally, dozens of computational tools have been developed to aid the process of variant pathogenicity identification.

Current SNV pathogenicity prediction tools assess various properties of genetic variants and provide a likelihood of causing a disease^1–10^. This information aids in variant prioritization – the process of narrowing down the list of potential pathogenic variants, and, therefore, facilitating diagnostics^11^. Variant pathogenicity prediction tools often utilize evolutionary conservation^7^, protein-level features^2^, and biochemical properties of amino acids^6^ to computationally predict the impact of a sequence change on protein structure and/or function. More recently, ensemble models have been developed, which combine pathogenicity scores from multiple models to generate a collective prediction^3–5,9,12^.

To assess the performance of SNV pathogenicity prediction tools, datasets of clinically classified variants, such as ClinVar^13^ and HGMD^14^, are commonly employed. These datasets contain large numbers of clinically relevant variants, making them indispensable in the development and validation of gene prioritization models. Despite that, previous benchmark studies showed that validation on such data yields an inflated performance estimate, that is not practically achieved for the variants not yet observed in ClinVar^15,16^.

In our study, we developed SNPred - an ensemble gradient boosting-based SNP pathogenicity prediction model. SNPred incorporates 33 pathogenicity prediction scores and 7 conservation scores. To train the model, we used a large dataset consisting of 229,336 variants obtained from ClinVar. SNPred was evaluated on the six different datasets comprising variants from saturation genome editing (SGE) study and ClinVar. It demonstrated superior performance compared to 32 other state-of-the-art SNV pathogenicity predictors, such as REVEL^3^, and BayesDel^9^, which were previously shown to outperform other meta-predictors on clinical data^17^.

During the validation process, we discovered several challenges that are inherent to validation of SNP pathogenicity predictors using ClinVar and may lead to inflated performance estimates. Firstly, we show that ClinVar variants on average tend to be more accurately classified than variants from a non-overlapping set obtained from SGE, as tested for *BRCA1*^18^. Secondly, we prove that the exceptionally high accuracy scores achieved by certain models on ClinVar are only attainable if the models overtrain by learning to misclassify variants that are already misclassified in ClinVar.

## Methods

### SNV pathogenicity prediction model

Figure 1 illustrates the outline of the final pipeline. To characterize each variant, we utilized 33 pathogenicity prediction scores, 7 conservation scores, and 42 gnomAD and EXaC allele frequencies from dbNSFP as features. For a comprehensive list of features and their corresponding importance, please refer to **Supplementary Table 1**. To efficiently gather this data, we employed the myvariant (v1.0.0) library in Python (v3.9.0). Subsequently, a gradient-boosting model was trained using these features. We considered three popular Python implementations, namely CatBoost, XGBoost, and LightGBM. To evaluate their performance, we employed six validation datasets, which consisted of five subsets of ClinVar and one BRCA1 SGE study dataset (**Figure 1B**). CatBoost achieved the highest performance among all models in four ClinVar datasets while securing the second-highest position in the remaining two datasets (**Figure S1 A-B**). As such, CatBoost v1.2 was used for the final ensemble model.

**Figure 1.**
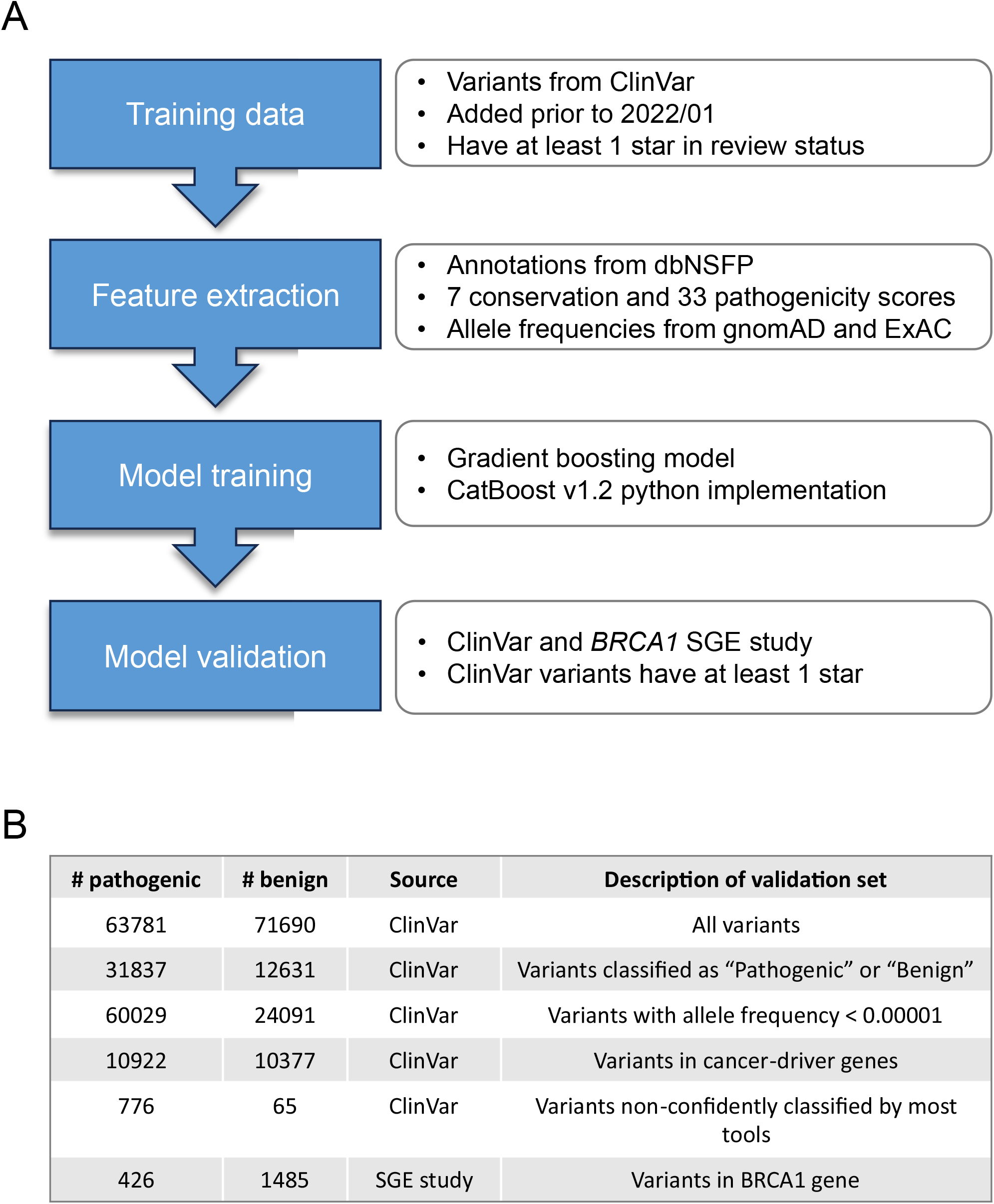
Training and validation of SNPred A: Details of the process of training and validation of SNPred B: Descriptions of six datasets used for validation of SNPred

We employed random search to investigate the model’s responsiveness to variations in hyperparameters from their default settings. These hyperparameters include the number and depth of trees, learning rate, L2 regularization, and the random strength (which determines the fraction of columns used when building each tree). For 100 combinations of hyperparameters for each dataset, we tested the resulting ROC AUC for two validation sets – variants from the *BRCA1* SGE study and all ClinVar variants that were added after January 2022, and have at least 1 star in assertion criteria. To avoid overtraining, our aim was not to select the best-performing parameters directly but rather to identify trends that consistently and robustly improved performance across the two different datasets. Among the various hyperparameters, the only change that consistently yielded better performance was an increase in L2 regularization (**Figure S1 C-F**). This finding is reasonable considering the utilization of highly correlated features, such as different pathogenicity prediction scores, allele frequencies in different populations, and various conservation scores.

### Usage of variants recently added to Clinvar to minimize overlap in training and validation

A validation dataset serves the purpose of evaluating a model’s performance on new and unseen data, providing an unbiased estimation of its ability to generalize. When the same data is used for both training and validation, the model may have already been exposed to and learned from that data during training, resulting in an inflated validation performance. This concern requires particular attention in the case of ensemble models, as the validation sets should have minimal overlap with the training sets of each base model used in the final ensemble.

While it is practically difficult to collect each base model’s training dataset, we can simply use only recently annotated variants found in databases. We looked at the changelog of dbNSFP to see that since April 6, 2021, there have been no changes to the pathogenicity prediction scores. As such, variants that were added to public databases after that point likely have not been used for training of any SNP pathogenicity prediction models. Because of that, for validation set assembly we used variants that are present in the latest versions of ClinVar (April 2023) but are absent in the older version (January 2022).

### Validation data

Six different validation datasets were used to assess the performance in different settings. The first five were taken from ClinVar (April 2023 version), where only variants with at least 1 ClinVar star that were added after January 2022 were considered. In the first dataset, no additional filtering was applied. In the second one, only variants that were classified as “Pathogenic” or “Benign” were used. The third dataset consisted of variants with AF < 0.00001. The fourth dataset contained variants in cancer driver genes from the Network of Cancer Genes^19^.

For the fifth dataset, we collected the variants from the SGE study of BRCA1. Variants that were classified as “intermediate” were filtered out – only “Functional” and “Non-functional” were used.

The sixth dataset comprised variants that lacked confident classification by the majority of tools. Specifically, we identified variants where only a percentage P of tools had a score higher than the Q quantile for pathogenic variants or lower than the (1-Q) quantile for benign variants. To ensure the reliability of these ambiguous variants in ClinVar, we included only those with at least two ClinVar stars in our selection. For the final validation set, we used P = 20% and Q = 0.8. However, we also examined the sensitivity of validation results by varying Q from 0.8 to 0.95 and P from 10% to 30% (**Figure S2**).

### Training data

For our analysis, we utilized variants classified as “Benign” (B), “Likely benign” (LB), “Benign / Likely benign,” “Pathogenic” (P), “Likely pathogenic” (LP), and “Pathogenic / Likely pathogenic” from the ClinVar database (as of April 2023). To ensure consistency, we only considered variants that were added to ClinVar before January 2022, which were obtained from the archived version of ClinVar from January 2022. Moreover, we specifically included variants with at least 1 review status star for training purposes.

## Results

### Performance comparison of SNPred to other predictors

To evaluate the performance of SNP pathogenicity models, we employed six validation datasets: five subsets of ClinVar and one *BRCA1* SGE study dataset (**Figure 1B**). The performance of SNPred was compared to 32 other variant pathogenicity predictors, for which the scores were taken from dbNSFP. For each validation set, we measured areas under the ROC and precision-recall curves (AUC ROC and AUC PR) to assess the quality of predictions, and Brier score^20^ to test the calibration of models (**Supplementary Tables 2-7**).

In all validations, SNPred showed superior performance in terms of AUC ROC (**Figure 2 A**). The particularly impressive marginal improvement over the second-best predictor was achieved for rare variants, variants in cancer-driver genes and variants non-confidently classified by most tools, where the difference in AUC ROC was 0.013, 0.015, and 0.033 respectively. The average gain in AUC ROC across all datasets over the second-best predictor, BayesDel, was 0.017. We also show that SNPred shows superior results for all levels of strictness when filtering ClinVar variants by assertion criteria stars (**Figure S3**).

**Figure 2.**
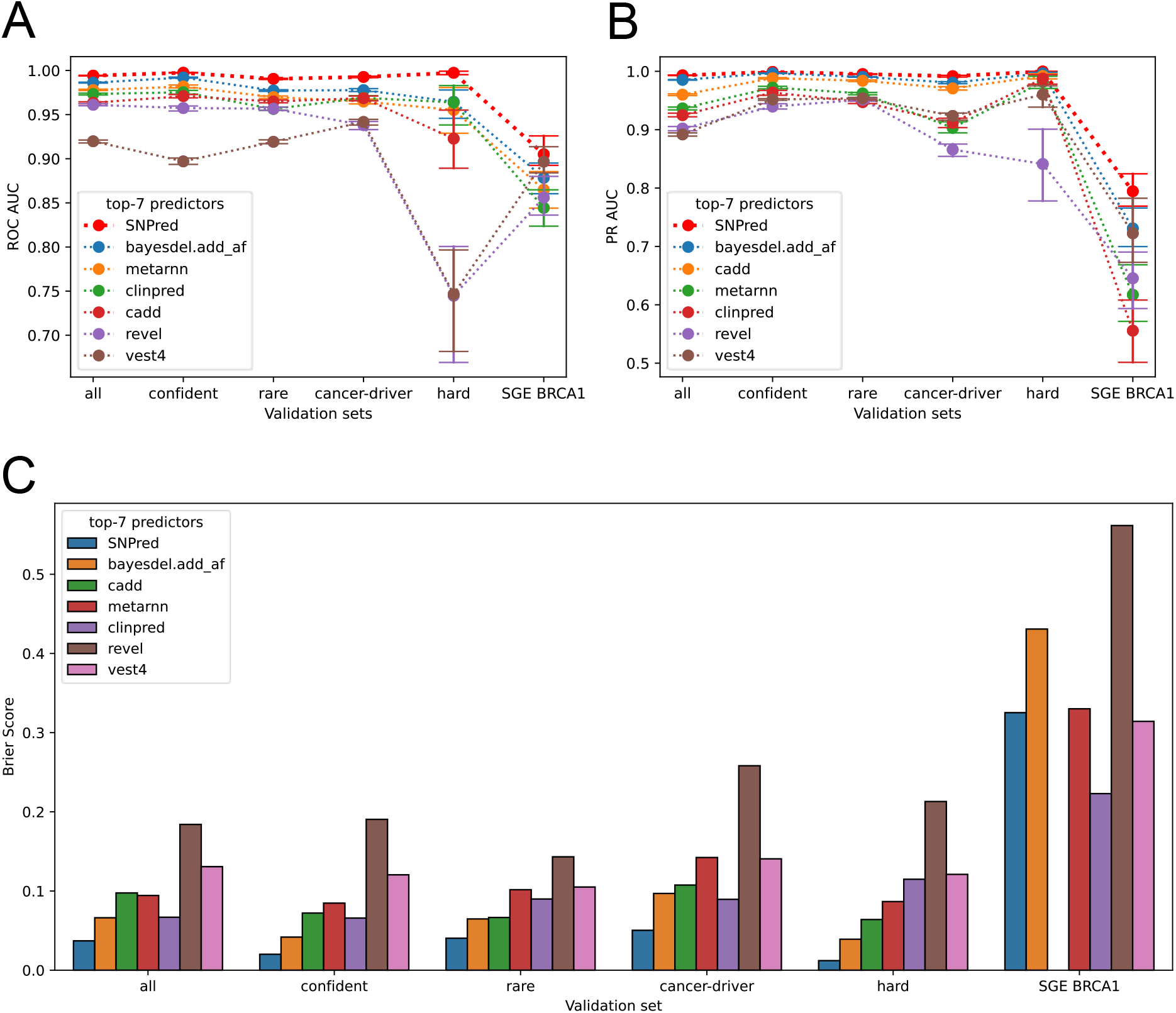
Comparison of SNPred’s performance to state-of-the-art SNP pathogenicity prediction tools. A: AUC ROC scores for the top-7 models across six validation datasets B: AUC PR scores for the top-7 models across six validation datasets C: Brier scores for the top-7 models across six validation datasets

The baseline area under the precision-recall curve is influenced by the class priors, making it difficult to compare the absolute values of AUC PR across different validation sets. However, it is possible to evaluate and compare the relative performance of models in terms of AUC PR for the same dataset. In this context, SNPred exhibited superior performance compared to other predictors across all six validation sets (**Figure 2B**). The difference in performance was particularly notable for the variants from the *BRCA1* SGE study, where SNPred achieved an AUC PR of 0.794, while the second-best predictor, BayesDel, attained an AUC PR of 0.731.

The Brier score can be used as a metric to evaluate the accuracy and calibration of probabilistic predictions. It measures the mean squared difference between predicted probabilities and the actual outcomes. The lower the Brier score, the better the model’s calibration and predictive accuracy. For all validation sets derived from ClinVar, SNPred showed the lowest Brier scores, which on average (geometric mean) were 1.9 times lower than that of the second-best predictor, BayesDel (**Figure 2 C**). However, for variants from the *BRCA1* SGE study, all predictors performed poorly in terms of Brier score. In this specific context, SNPred did not demonstrate the best results – among the top-7 best performing predictors, ClinPred^4^ and Vest4^10^ showed lower Brier scores. The very poor Brier score of all models can be explained by the generally high pathogenicity scores assigned to the benign variants from the *BRCA1* SGE study (**Figure S4 A**,**B**). This poor calibration of models also poses a challenge in choosing a threshold that discriminates benign and pathogenic variants, because the same threshold corresponds to vastly different precision values for ClinVar and SGE variants (**Figure S4 C**,**D**).

### BRCA1 variants from ClinVar are classified significantly better than those from SGE

In several published benchmarking studies, it was noticed that performance metrics estimated using ClinVar are higher than those on variants that are not yet observed in ClinVar^15,16^. However, these studies often employ datasets that significantly differ from each other, making it challenging to attribute the performance discrepancy solely to an inherent issue with ClinVar. Other factors such as variations in the diseases of interest, distribution of occurrences of altered genes, and other dataset characteristics may also contribute to the observed differences in performance estimation.

Here, we aimed to check whether the performance estimate on ClinVar would still be inflated if the discussed above confounding factors were controlled for. To do that, we gathered two non-intersecting datasets: (i) all *BRCA1* variants with at least 1 review status star that were added to ClinVar after January 2022, and (ii) variants from the *BRCA1* SGE study that are not in ClinVar. Then, we took 12 well-performing predictors, that have AUC ROC of at least 0.8 for both datasets and compared their AUC ROC on these two datasets. 12 out of 12 predictors showed higher performance on ClinVar variants (**Figure 3 A**), and on average all predictors showed an increase in AUC ROC of 0.056 in ClinVar compared to SGE data.

**Figure 3.**
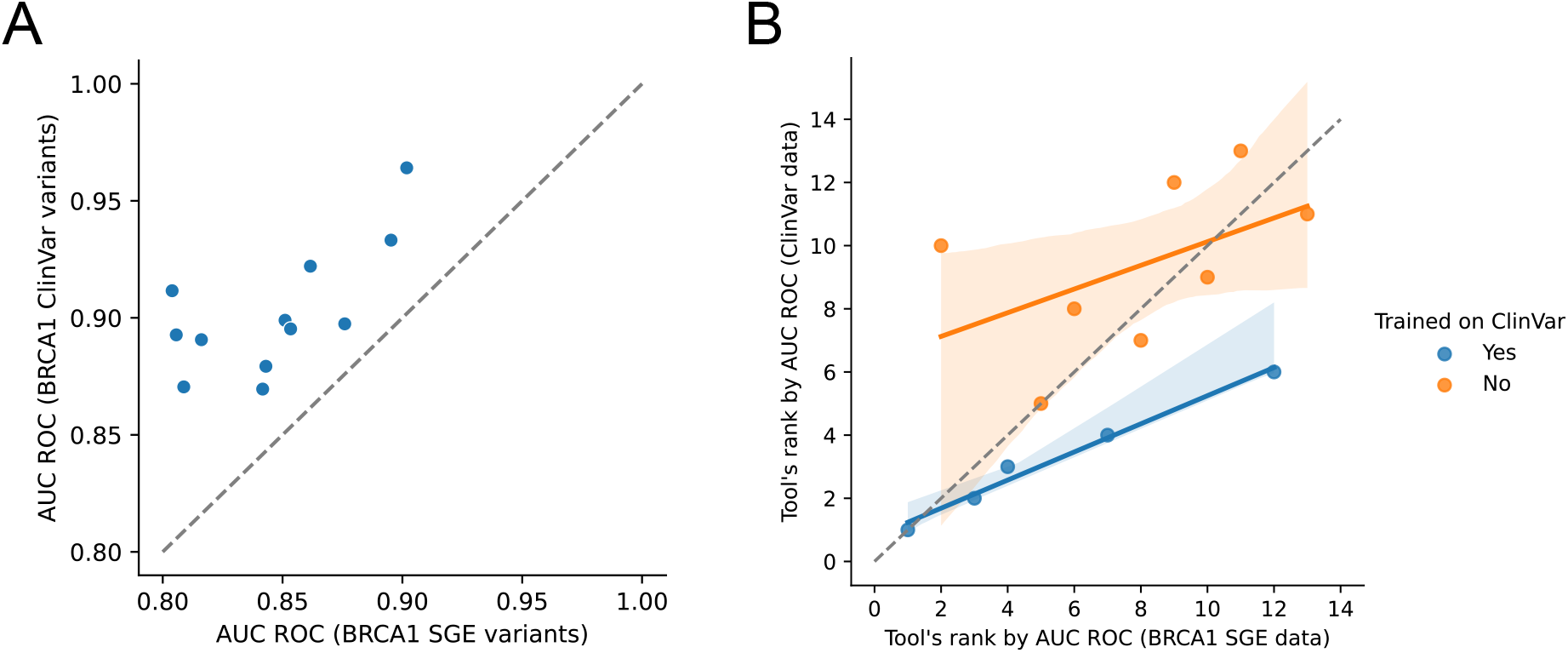
Comparison of SNP pathogenicity predictors’ performance on ClinVar and SGE variants A: Scatter plot of AUC ROC scores for 12 predictions on *BRCA1* variants from ClinVar and *BRCA1* SGE study B: Relative performance of predictors on ClinVar and *BRCA1* SGE variants

This finding corroborates the hypothesis that ClinVar variants in general are easier to classify using computational tools. One explanation for this is that easily classified variants are discovered and submitted to ClinVar more frequently, making them overrepresented in the database.

### SNP pathogenicity prediction models tend to replicate ClinVar misclassifications

Genetic analysts typically determine whether a variant is (likely) pathogenic or benign using standardized criteria such as those defined by the American College of Medical Genetics^21^. Information from variant annotations, genetic variation databases, computational tools, and other sources is collectively evaluated as evidence for or against pathogenicity. The ultimate classification of a variant under such criteria may be submitted to public databases such as ClinVar to further inform future classifications of the same variant or variants in the same gene.

The ACMG guidelines acknowledge the potential for mistakes. For instance, terms like “likely pathogenic” or “likely benign” indicate a confidence level of over 90% that a variant causes a disease or is benign^21^. ClinVar also has a lot of old classifications, such as those pulled in from the OMIM database which can be years or decades out of date. Many variants are later reclassified downward based on experimental or population allele frequency information which was not available to the genetic analyst at the time they interpreted the variant. This means that errors in classification are to be expected in databases like ClinVar. However, when a dataset containing inaccuracies is used for evaluation, predictive models can only achieve the highest possible performance scores if they replicate those incorrect classifications.

Formally, a model learns to replicate misclassifications if *P*(*Y*_*model*_ | *Y*_*true*_,*Y*_*ClinVar*_) is not equal to *P*(*Y*_*model*_ | *Y*_*true*_). In this context, we investigated whether the top-performing models’ high accuracy scores on ClinVar can be achieved without imitating ClinVar’s misclassifications. This essentially means examining if the condition *P*(*Y*_*model*_ | *Y*_*true*_,*Y*_*ClinVar*_) = *P*(*Y*_*model*_ | *Y*_*true*_) is satisfied. Equivalently, it means that

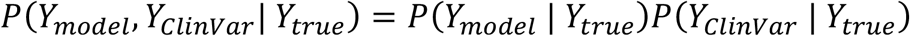

Under the assumption of conditional independence, the accuracy can be computed from the True Positive Rate (TPR) and True Negative Rate (TNR) of the SNP pathogenicity prediction model and ClinVar by the formula:

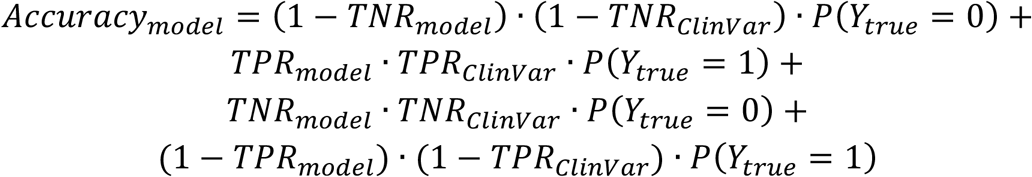

A more detailed proof of the formula is provided in the **Supplementary Materials**. Several implications follow from the formula. The observed accuracy of SNPred on the recently added ClinVar variants, 65% of which are Classified as “Likely pathogenic” or “Likely benign”, is 0.968. If we assume that the model is not more accurate than ClinVar, to get the observed SNPred accuracy of 0.968 (when using all variants added to ClinVar after January 2022) the TPR and TNR of both ClinVar and the model must be higher than 98.4%. For a perfect model to achieve such observed accuracy, the accuracy of ClinVar itself should be at least 96.8%.

Equivalently, if a model has a misclassification rate that is the same as or higher than ClinVar’s misclassification rate, and it manages to reach the observed accuracy of 0.968, then ClinVar’s own misclassification rate must be lower than 1.6%. Furthermore, even if we were to consider a perfectly accurate model, ClinVar’s misclassification rate would still need to be under 3.2% to attain the same level of observed accuracy.

While it is hard to accurately calculate the rate of variant misclassification in ClinVar, it is possible to roughly estimate it. One study that focused on reclassification of variants in ClinVar found that around 20% of “Likely pathogenic” and around 7% of “Likely benign” variants were downgraded after reclassification^22^. Another reanalysis of variants with AF > 0.005 in at least one gnomAD ancestry that are classified as pathogenic in ClinVar, found that 40% of variants should be downgraded to “Benign” / “Likely benign” / “Unknown significance”^23^. Another study concluded that of all the variants classified as P/LP that were reclassified from 2014 to 2020, 8% were downgraded^24^. Analysis of concordance between different submitters has found that only 89.3% of variants had a majority (> 66%) agreement about pathogenicity status^25^. For the *BRCA1* gene, 95.9% of variants classified as pathogenic and 95.5% of variants classified as benign in ClinVar were successfully replicated using SGE^18^. Out of 1352 P/LP ClinVar variants found in Deciphering Developmental Disorder (DDD) study probands, 106 were found to be benign – TPR = 1246/1352 = 92.1%^26^.

Therefore, according to the existing literature, the assumptions we made in our analysis – using ClinVar’s misclassification rates of at least 1.6% or 3.2% – can be considered conservative. Consequently, it can be inferred that the advanced models currently available cannot attain the observed accuracy of 0.968 or higher without emulating the misclassifications identified in ClinVar.

### Predictors that are trained on ClinVar have higher relative performance on ClinVar compared to SGE

We hypothesized that models that use ClinVar in their training data may be excessively overtrained to ClinVar. In such case, these models should also better replicate the incorrect classifications present in ClinVar data. As a result, this could potentially cause additional inflation in performance estimates when these models are evaluated on ClinVar data.

We compared SNV pathogenicity predictors using both ClinVar and SGE data. Only predictors that had AUC ROC of at least 0.8 for both ClinVar and SGE data were used, and only variants added to ClinVar after January 2022 were selected to ensure they did not overlap with ClinVar data used for model training. We then compared how predictors were ranked on each of the two datasets (**Figure 3 B**). Every model that used ClinVar for training was ranked the same or higher in validation involving ClinVar data compared to SGE data. The difference in relative performance was more significant (Mann-Whitney U-test p-value = 0.037) for models that were trained using ClinVar than for those that were not.

It’s important to note that further investigation with more diverse sets of data is necessary to establish a definitive conclusion. However, our findings strongly indicate that using ClinVar for performance estimation could potentially lead to inflated performance assessments for models trained on ClinVar, even if the ClinVar data used for training and validation do not overlap.

## Discussion

We developed SNPred, an ensemble gradient boosting model for predicting the pathogenicity of nonsynonymous single nucleotide variants (nsSNVs). SNPred uses 7 conservation scores, 33 pathogenicity prediction scores, and allele frequencies from ExAC and gnomAD to create an aggregate pathogenicity score. The resulting scores were precalculated for all possible nsSNVs and were provided for public use.

One of the main strengths of the SNPred is the utilization of a higher number of pathogenicity scores. While such models as BayesDel^9^, REVEL^3^, MetaRNN^5^, and ClinPred^4^ only use 7-18 pathogenicity scores as features, SNPred utilizes 33 scores. Additionally, SNPred was trained on more recent ClinVar variants, which tend to be classified more accurately than the older ones ^24^.

SNPred’s performance was assessed using six distinct validation datasets obtained from ClinVar and *BRCA1* saturation genome editing data. It consistently outperformed other state-of-the-art tools, including REVEL, ClinPred, BayesDel, and MetaRNN, particularly excelling with rare and cancer-related variants. Importantly, this performance difference remained consistent regardless of the assertion criteria filtering options applied to ClinVar. This finding indicates that the disparity in performance is unlikely to be attributed to false positive data in ClinVar^16^ but rather to the genuinely improved capabilities of SNPred.

However, in these validations, a few other predictors besides SNPred, such as BayesDel and MetaRNN, performed relatively well, even if they were inferior to SNPred. Thus, to compare the tools on variants that specifically pose a significant challenge to computational predictors, we assembled a dataset of variants that are not confidently classified by the majority of tools but are reliably classified in ClinVar. For these variants, SNPred showed by far the best performance, outperforming the next four leading predictors by a margin of 0.03 to 0.08. This shows that SNPred can confidently classify variants that were ambiguously classified by the previously available computational tools.

During the validation process, we discovered several challenges that are inherent to validation of SNP pathogenicity predictors using ClinVar. These challenges could potentially result in inaccurate assessments of performance.

First, by making specific assumptions about the misclassification rate in ClinVar, we demonstrate that the remarkably high accuracy scores obtained by certain models on ClinVar can only be achieved through overtraining, wherein the models learn to misclassify variants that are already misclassified in ClinVar. To validate these assumptions, we conducted a thorough analysis of the literature on misclassification rates and found that the misclassification rate in ClinVar consistently surpasses the lower bound assumed in our analysis. Because models learn to replicate misclassification in ClinVar, the comparison of performance using ClinVar data may not be adequate for predictors with very high accuracy, because it is hard to distinguish between a predictor that genuinely classifies variants more accurately and a predictor that is able to replicate misclassifications more effectively. To alleviate this problem, it might be useful to apply stricter filtering on the assertion criteria, thus decreasing the number of false positive data points.

Next, we sought to empirically examine the hypothesis that variants that are accurately classified using computational tools appear in ClinVar more frequently, making them overrepresented in the database. To that end, we considered a set of *BRCA1* variants from ClinVar and a non-intersecting set of *BRCA1* variants from an SGE study. We tested how 12 top-performing predictors would perform on these two datasets. Confirming our hypothesis, all of the predictors classified ClinVar variants more accurately, with the mean difference in AUC ROC of 0.056 between the two datasets. This problem is much harder to get around through filtering and is inherent to databases such as ClinVar. It indicates that validation of any pathogenicity prediction algorithm using ClinVar data may result in inflated performance estimates. Therefore, to ensure reliable validation, we recommend using ClinVar variants that have good assertion criteria, as well as other sources of data, such as SGE studies^18,27,28^.

## Data Availability

SNPred pathogenicity scores for all possible non-synonymous changes in human genome can be found at https://www.synapse.org/#!Synapse:syn52137034/files/.
The source code to run SNPred is available at: https://github.com/ArtomovLab/SNPred

https://www.synapse.org/#!Synapse:syn52137034/files/

## Data availability

SNPred pathogenicity scores for all possible non-synonymous changes in the human genome can be found at https://www.synapse.org/#!Synapse:syn52137034/files/. The source code to run SNPred is available at: https://github.com/ArtomovLab/SNPred

## Acknowledgements

The authors would like to thank the Institute for Genomic Medicine (Nationwide Children’s Hospital) community for providing insightful comments and support.

## References

1. Rentzsch, P., Witten, D., Cooper, G. M., Shendure, J. & Kircher, M. CADD: Predicting the deleteriousness of variants throughout the human genome. Nucleic Acids Res 47, D886–D894 (2019).

2. Adzhubei, I. A. et al. A method and server for predicting damaging missense mutations. Nature Methods vol. 7 248–249 Preprint at 10.1038/nmeth0410-248 (2010).

3. Ioannidis, N. M. et al. REVEL: An Ensemble Method for Predicting the Pathogenicity of Rare Missense Variants. Am J Hum Genet 99, 877–885 (2016).

4. Alirezaie, N., Kernohan, K. D., Hartley, T., Majewski, J. & Hocking, T. D. ClinPred: Prediction Tool to Identify Disease-Relevant Nonsynonymous Single-Nucleotide Variants. Am J Hum Genet 103, 474–483 (2018).

5. Li, C., Zhi, D., Wang, K. & Liu, X. MetaRNN: differentiating rare pathogenic and rare benign missense SNVs and InDels using deep learning. Genome Med 14, (2022).

6. Niroula, A., Urolagin, S. & Vihinen, M. PON-P2: Prediction method for fast and reliable identification of harmful variants. PLoS One 10, (2015).

7. Sim, N. L. et al. SIFT web server: Predicting effects of amino acid substitutions on proteins. Nucleic Acids Res 40, (2012).

8. Frazer, J. et al. Disease variant prediction with deep generative models of evolutionary data. Nature 599, 91–95 (2021).

9. Feng, B. J. PERCH: A Unified Framework for Disease Gene Prioritization. Hum Mutat 38, 243–251 (2017).

10. Carter, H., Douville, C., Stenson, P. D., Cooper, D. N. & Karchin, R. Identifying Mendelian disease genes with the variant effect scoring tool. BMC Genomics 14 Suppl 3, (2013).

11. Eilbeck, K., Quinlan, A. & Yandell, M. Settling the score: Variant prioritization and Mendelian disease. Nature Reviews Genetics vol. 18 599–612 Preprint at 10.1038/nrg.2017.52 (2017).

12. Dong, C. et al. Comparison and integration of deleteriousness prediction methods for nonsynonymous SNVs in whole exome sequencing studies. Hum Mol Genet 24, 2125–2137 (2015).

13. Landrum, M. J. et al. ClinVar: Public archive of interpretations of clinically relevant variants. Nucleic Acids Res 44, D862–D868 (2016).

14. Stenson, P. D. et al. The Human Gene Mutation Database (HGMD®): optimizing its use in a clinical diagnostic or research setting. Human Genetics vol. 139 1197–1207 Preprint at 10.1007/s00439-020-02199-3 (2020).

15. Gunning, A. C. et al. Assessing performance of pathogenicity predictors using clinically relevant variant datasets. J Med Genet 58, 547–555 (2021).

16. Li, J. et al. Performance evaluation of pathogenicity-computation methods for missense variants. Nucleic Acids Res 46, 7793–7804 (2018).

17. Tian, Y. et al. REVEL and BayesDel outperform other in silico meta-predictors for clinical variant classification. Sci Rep 9, (2019).

18. Findlay, G. M. et al. Accurate classification of BRCA1 variants with saturation genome editing. Nature 562, 217–222 (2018).

19. Repana, D. et al. The Network of Cancer Genes (NCG): A comprehensive catalogue of known and candidate cancer genes from cancer sequencing screens. Genome Biol 20, (2019).

20. Rufibach, K. Use of Brier score to assess binary predictions. Journal of Clinical Epidemiology vol. 63 938–939 Preprint at 10.1016/j.jclinepi.2009.11.009 (2010).

21. Richards, S. et al. Standards and guidelines for the interpretation of sequence variants: A joint consensus recommendation of the American College of Medical Genetics and Genomics and the Association for Molecular Pathology. Genetics in Medicine 17, 405–424 (2015).

22. Harrison, S. M. & Rehm, H. L. Is ‘likely pathogenic’ really 90% likely? Reclassification data in ClinVar. Genome Medicine vol. 11 Preprint at 10.1186/s13073-019-0688-9 (2019).

23. Xiang, J. et al. Reinterpretation of common pathogenic variants in ClinVar revealed a high proportion of downgrades. Sci Rep 10, (2020).

24. Sharo, A. G., Zou, Y., Adhikari, A. N. & Brenner, S. E. ClinVar and HGMD genomic variant classification accuracy has improved over time, as measured by implied disease burden. Genome Med 15, 51 (2023).

25. Yang, S. et al. Sources of discordance among germ-line variant classifications in ClinVar. Genetics in Medicine 19, 1118–1126 (2017).

26. Wright, C. F. et al. Evaluating variants classified as pathogenic in ClinVar in the DDD Study. doi:10.1038/s41436.

27. Meitlis, I. et al. Multiplexed Functional Assessment of Genetic Variants in CARD11. Am J Hum Genet 107, 1029–1043 (2020).

28. Mhl, A. & Jrb, P. Saturation genome editing of DDX3X clarifies pathogenicity of germline and somatic variation. doi:10.1101/2022.06.10.22276179.

