## Supplementary materials for "SNPred outperforms other ensemble-based SNV pathogenicity predictors and elucidates the challenges of using ClinVar for evaluation of variant classification quality"

Ivan Molotkov[1,2], Daniel C. Koboldt[1,2], Mykyta Artomov[1,2]

1 – The Steve and Cindy Rasmussen Institute for Genomic Medicine, Nationwide Children's Hospital, Columbus, OH, 43215

2 – Department of Pediatrics, The Ohio State University College of Medicine, Columbus, OH, 43205

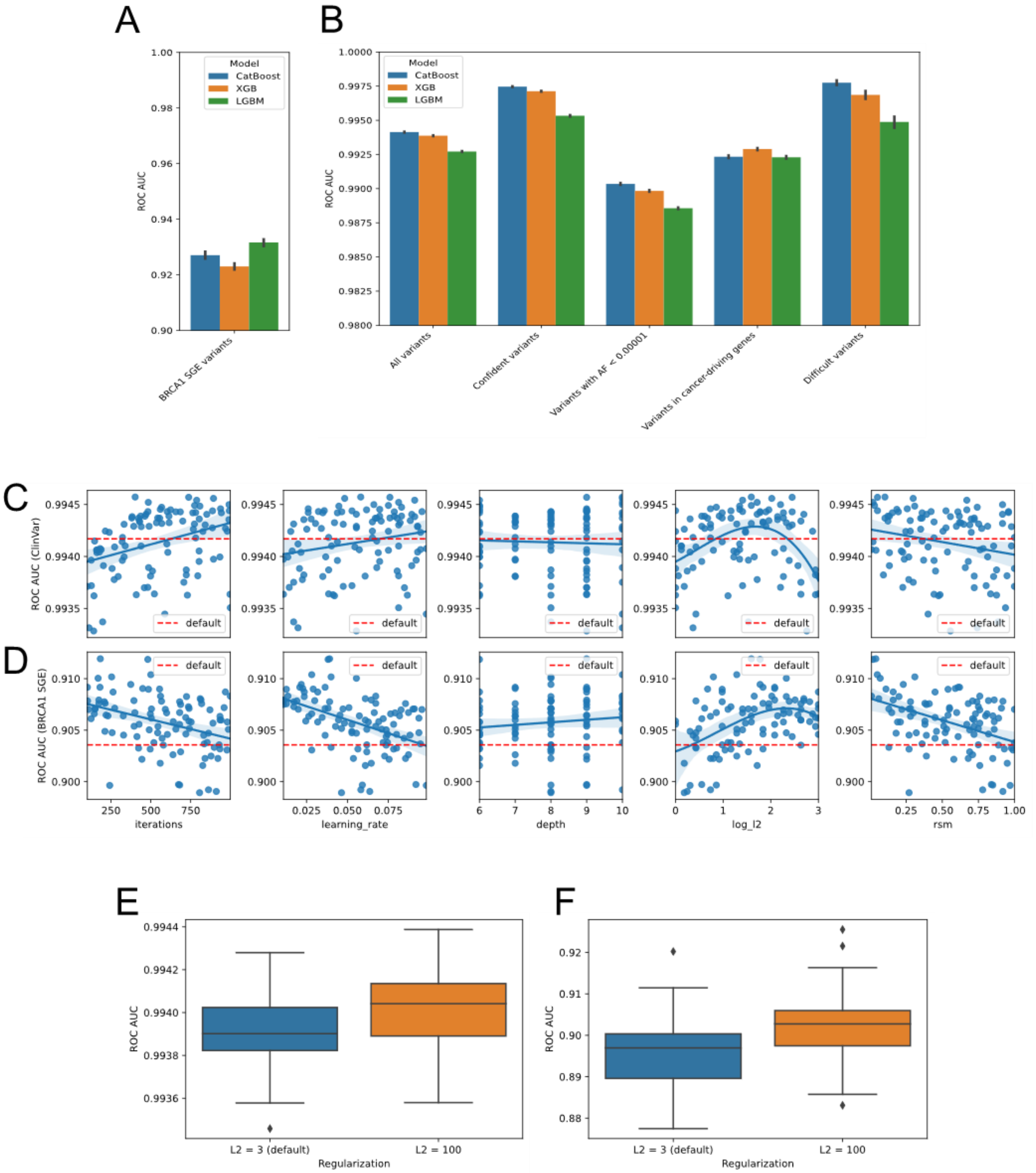


**Figure S1**. Model selection process for SNPred

A-B: Comparison of ROC AUC of CatBoost, XGBoost and LightGBM on six validation sets.

C-D: Relation between CatBoost hyperparameter selection and ROC AUC score on ClinVar (C) and *BRCA1* SGE variants (D)

E-F: Bootstrapped ROC AUC scores of CatBoost with default and strong L2 regularization for ClinVar (E) and *BRCA1* SGE variants (F)

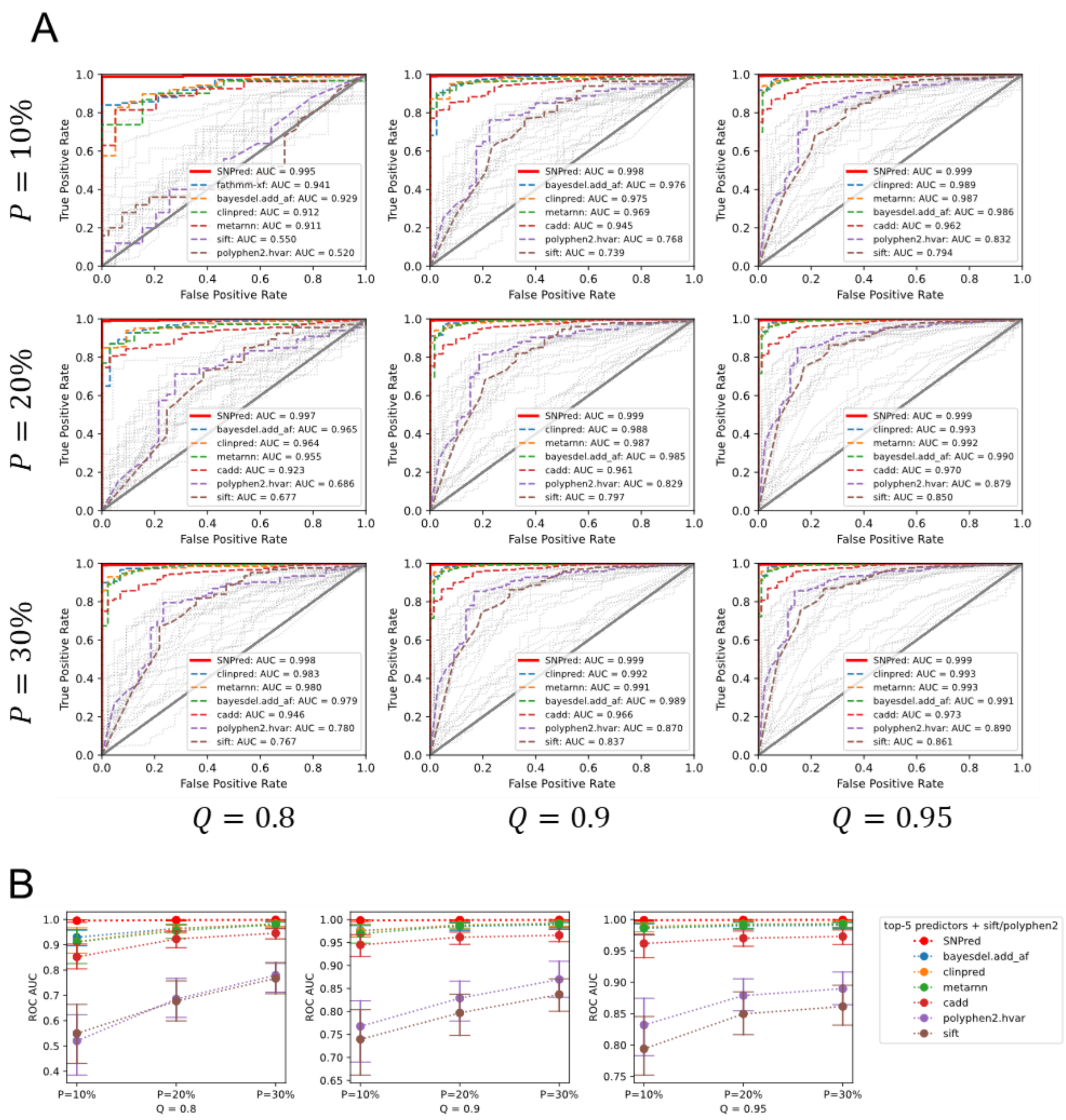


**Figure S2**. Performance on variants where only a percentage P of tools had a score higher than the Q quantile for pathogenic variants or lower than the (1-Q) quantile for benign variants.

A: ROC-curves for P ranging from 10% to 30%, Q – from 0.8 to 0.95.

B: AUC ROC for P ranging from 10% to 30%, Q – from 0.8 to 0.95.

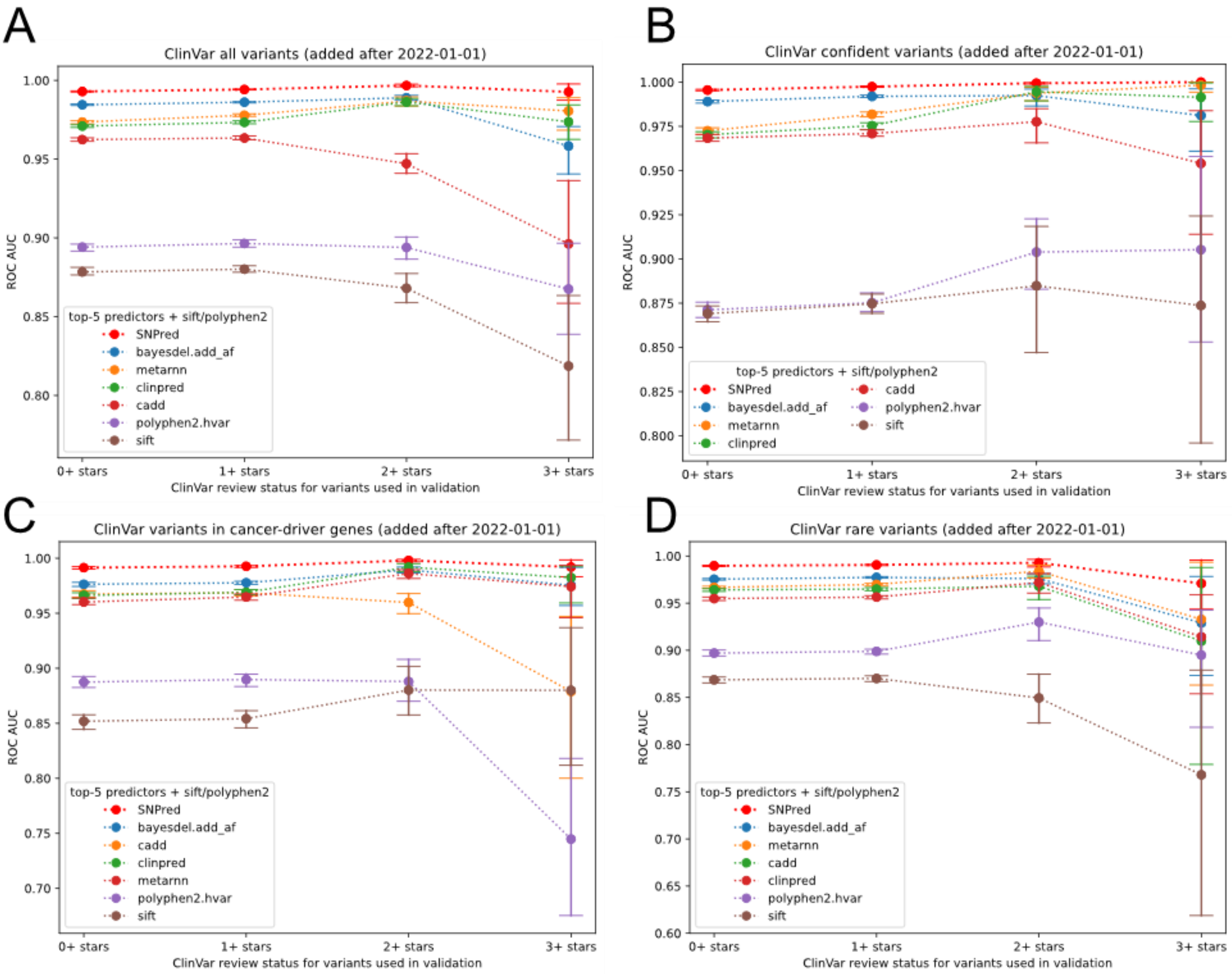


**Figure S3**. Comparison of performance using ClinVar variants across four different datasets after applying varying levels of strictness on the review status.

A: All ClinVar variants added after January 2022

B: Variants that are classified as “Pathogenic” or “Benign” in ClinVar

C: ClinVar variants in cancer-driver genes

D: ClinVar variants with allele frequency less than 0.00001

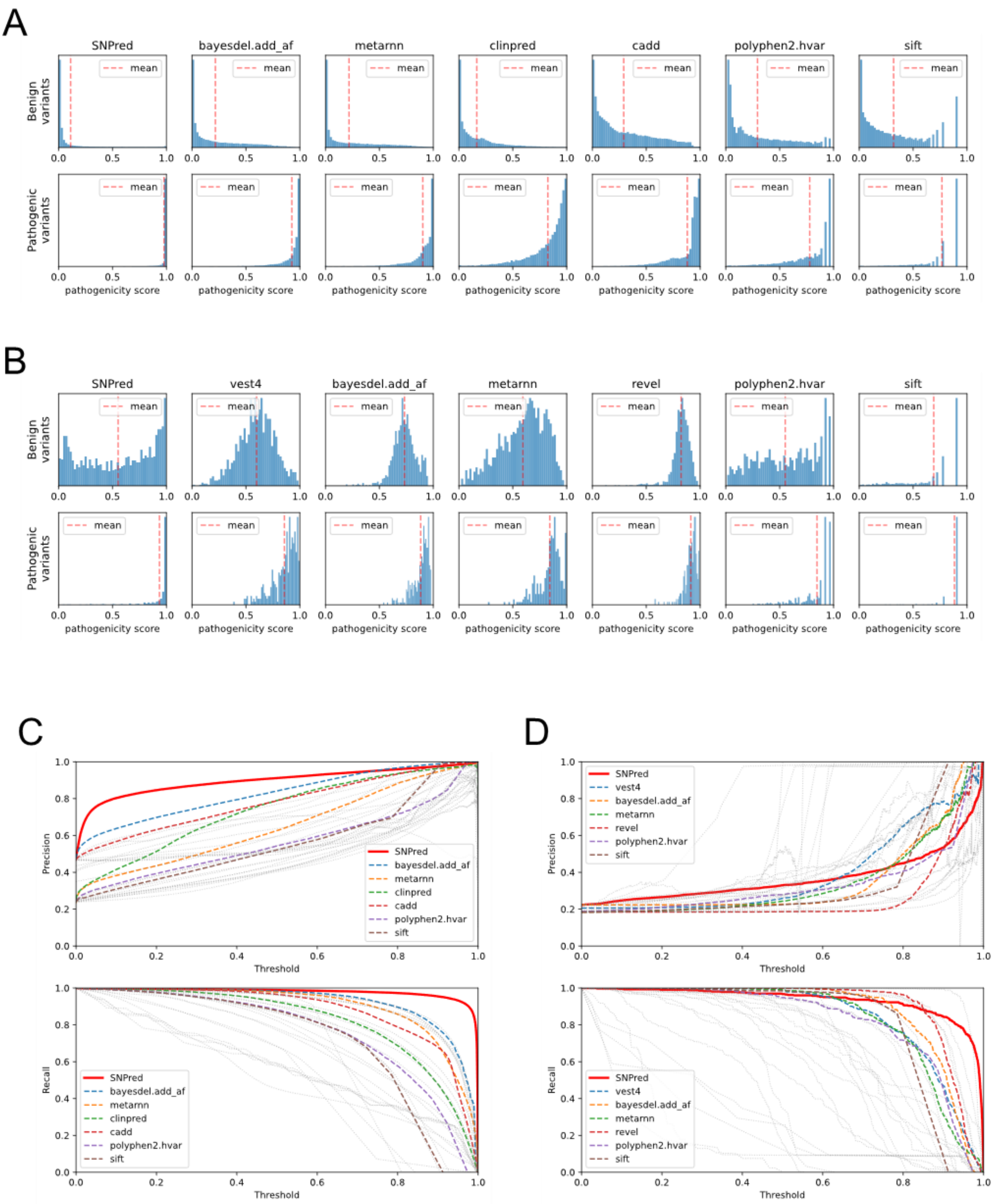


**Figure S4**. Inflated pathogenicity scores for benign variants in *BRCA1* lead to low precision for most threshold selections.

A-B: Distribution of scores for 5 best predictors and SIFT, PolyPhen2 on benign and pathogenic variants from ClinVar (C) and *BRCA1* SGE study (D)

C-D: Precision and recall curves obtained for ClinVar variants (A) and *BRCA1* SGE variants (B)

*Upper bound on accuracy achievable by a model without replicating ClinVar's misclassifications*

If a model tends to misclassify a variant when it is already misclassified in ClinVar, it indicates that the model is learning to replicate ClinVar misclassifications. In order to avoid this, it is necessary for the classifications made by ClinVar and the model to be independent of each other, considering the true classification of the variants. Then, the estimated accuracy of a model can be expressed using True Positive Rate and True Negative Rate values for ClinVar's and the model's classifications as follows:

$$
\begin{gathered}
Accuracy_{model} = \\
P(Y_{model} = 1, Y_{ClinVar} = 1, Y_{true} = 0) + \\
P(Y_{model} = 1, Y_{ClinVar} = 1, Y_{true} = 1) + \\
P(Y_{model} = 0, Y_{ClinVar} = 0, Y_{true} = 0) + \\
P(Y_{model} = 0, Y_{ClinVar} = 0, Y_{true} = 1) = \\
P(Y_{model} = 1 \mid Y_{true} = 0) P(Y_{ClinVar} = 1 \mid Y_{true} = 0) P(Y_{true} = 0) + \\
P(Y_{model} = 1 \mid Y_{true} = 1) P(Y_{ClinVar} = 1 \mid Y_{true} = 1) P(Y_{true} = 1) + \\
P(Y_{model} = 0 \mid Y_{true} = 0) P(Y_{ClinVar} = 0 \mid Y_{true} = 0) P(Y_{true} = 0) + \\
P(Y_{model} = 0 \mid Y_{true} = 1) P(Y_{ClinVar} = 0 \mid Y_{true} = 1) P(Y_{true} = 1) = \\
FPR_{model} \cdot FPR_{ClinVar} \cdot P(Y_{true} = 0) + \\
TPR_{model} \cdot TPR_{ClinVar} \cdot P(Y_{true} = 1) + \\
TNR_{model} \cdot TNR_{ClinVar} \cdot P(Y_{true} = 0) + \\
FNR_{model} \cdot FNR_{ClinVar} \cdot P(Y_{true} = 1) = \\
(1 - TNR_{model}) \cdot (1 - TNR_{ClinVar}) \cdot P(Y_{true} = 0) + \\
TPR_{model} \cdot TPR_{ClinVar} \cdot P(Y_{true} = 1) + \\
TNR_{model} \cdot TNR_{ClinVar} \cdot P(Y_{true} = 0) + \\
(1 - TPR_{model}) \cdot (1 - TPR_{ClinVar}) \cdot P(Y_{true} = 1)
\end{gathered}
$$

The first equation follows form the fact that model's classification is considered accurate if it matches ClinVar's classification. The second – from the assumption of conditional independence of ClinVar's and model's classifications given the true class. The last two equations follow from the definitions of TPR, TNR, FPR, FNR.

This formula allows to check the possible range of estimated accuracy of a model given TPR/TNR of ClinVar's classifications.

Under the assumption that TPR/TNR of ClinVar is greater than 0.5, the maximum accuracy will be achieved by a model with TPR and TNR of 1.0 (Accuracy increases as we increase TPR/TNR). The resulting accuracy will equal to $TPR_{ClinVar} \cdot P(Y_{true} = 1) + TNR_{model} \cdot TNR_{ClinVar} \cdot P(Y_{true} = 0)$. Thus, the estimated accuracy cannot be greater than the weighted average of TPR and TNR of ClinVar's classifications.
